# An effective volunteer community-based COVID-19 response program: the Vashon, WA Medical Reserve Corp Experience

**DOI:** 10.1101/2022.08.26.22279283

**Authors:** James Bristow, Clayton Olney, Vashon MRC COVID-19 Steering Committee, John Weinshel, Robert Rovig, Rick Wallace, Karla J. Lindquist

## Abstract

**Background:** Vashon, WA is a rural community at risk from COVID-19 due to advanced age and limited access to acute care. Medical Reserve Corps are a national network of 800 volunteer healthcare organizations that have contributed to the pandemic response in many communities. Here we evaluate the effectiveness of the Vashon Medical Reserve Corp’s (VMRC) volunteer, community-based COVID-19 response program that integrated public engagement, SARS-CoV2 testing, contact tracing, vaccination and material support in reducing COVID-19 transmission and severe disease.

**Methods:** This observational cross-sectional study compares cumulative COVID-19 case, hospitalization and death rates on Vashon with other King County zip codes and the county at large from February 2020 through November 2021. We developed multiple linear regression models of COVID-19 rates using metrics of age, race/ethnicity, wealth and educational attainment across King County zip codes. Effectiveness of contact tracing was evaluated by timeliness and success of case investigations, and identification and testing of named contacts. Vaccination effectiveness was estimated by comparing time to reach vaccination milestones. We examined vehicle traffic on Vashon ferries and King County highways to understand whether reduced mobility contributed to Vashon’s reduced COVID-19 rates.

**Results:** Vashon’s cumulative COVID-19 case rate was 29% that of King County overall and was lower across all age groups and races/ethnicities (both p<.01). A multiple linear regression model showed Vashon to be a significant outlier among King County zip codes with an observed rate 38% of predicted (p<.05), the lowest of any King County zip code. Vashon’s observed COVID-19 hospitalization and death rates were 22% and 32% of those predicted by parallel regression models. Hence, Vashon’s demographics do not explain its reduced COVID rates. Traffic reductions on King County highways and Vashon ferries were nearly identical throughout the study period suggesting altered mobility also does not explain Vashon’s low COVID-19 rates. Effectiveness of VMRC’s COVID-19 response program was demonstrated by 1) highly effective contact tracing that rapidly interviewed 93% of cases and subsequently tested 96% of named contacts, and 2) attainment of vaccination milestones 1-4 months earlier than comparable King County zip codes (p<.01).

**Conclusion:** VMRC’s volunteer, COVID-19 response program was associated with significantly fewer COVID-19 cases than predicted from its demographics. VMRC’s contact tracing and vaccination efforts were highly successful and likely contributed to reduced COVID-19 rates. The VMRC experience suggests that a decentralized community-based public health program can be highly effective in implementing epidemic control strategies when focused on an at-risk community. We suggest that MRCs can be particularly effective in extending the reach of county public health departments and should be included in ongoing pandemic planning.

## Introduction

Vashon WA, an island community of 11,000 people located in unincorporated King County, near metropolitan Seattle, shares increased health risks from COVID-19 with other rural US communities-an aging population with attendant comorbidities, no immediate access to acute care and a Federally Qualified Health Clinic that is under chronic financial stress (1,2). Vashon’s risk is compounded by a recent history of significant vaccine hesitancy (3).

Medical Reserve Corps (MRCs) are a network of ∼800 volunteer medical organizations operating under the auspices of the Assistant Secretary for Preparedness and Response, in the U.S. Department of Health and Human Services, with the goal of strengthening public health, improving emergency response capabilities, and building community resiliency (4). In March, 2020 as Seattle became the nation’s first SARS-CoV2 “hotspot’ (5), the Vashon Medical Reserve Corps (VMRC) and VashonBePrepared, two community-based volunteer emergency preparedness organizations, established an integrated COVID-19 response program with the specific goal of reducing SARS-CoV2 transmission in the community. This integrated response consisted of regular community engagement and education, free SARS-CoV2 testing, rapid case investigation/contact tracing, an aggressive vaccination campaign and material support for effected families. In this report we examine the contributions of Vashon’s demography, its COVID-19 response program, and its geography to its low COVID-19 rates.

## Materials and Methods

### Vashon’s COVID-19 Public Health Response

The Vashon COVID-19 response program was comprised of 3 primary efforts-community engagement, community health and community support and was organized into a typical incident command structure led by an emergency operations center (Fig S1). VMRC coordinated with Public Health-Seattle King County (PHSKC), but operated independently of it.

The primary tool for community engagement tool was twice weekly “[situation reports](http://vashonbeprepared.org/News/SitReps)” published in the local newspaper and emailed in English or Spanish to >3500 residents. VMRC operated a COVID-19 hotline 50 hours/week to provide public health guidance and schedule SARS-CoV2 tests. VMRC also advised more than 40 local businesses and schools about COVID-19 controls or management of outbreaks.

During the study period VMRC collected 5,716 COVID-19 samples at a drive-through site using supervised patient-collected nasal swab specimens (6) to minimize volunteer exposure and the amount of personal protective equipment required. SARS-CoV2 PCR was carried out by a CLIA approved, CAP accredited laboratory (Atlas, Genomics, Seattle WA). Results were usually returned to VMRC within 24 hours and always within 48 hours. The demographics of VMRC testing closely mirrored Vashon’s population (S2 Table).

VMRC investigated 214 cases (89% of all Vashon cases) during the study period. A team of four physicians delivered positive VMRC test results directly to patients by telephone, always within 24 hours of test completion, and then conducted interviews to identify contacts and provide isolation and quarantine advice. From Jan-September 2021, PHSKC referred Vashon cases to VMRC for contact tracing. In July 2021, VMRC’s contact tracing protocol was streamlined to focus on household contacts who comprised >80% of infected contacts through June 2021.

Working with Vashon’s independent pharmacy, VMRC also delivered 17,013 doses of SARS-CoV2 vaccines, primarily at a drive through site (March-May 2021) and indoor vaccine clinics (October-November 2021).

During the study period, VashonBePrepared’s COVID Relief Fund distributed $546,000, primarily to local charities, and was supported by individual charitable donations and FEMA reimbursement for emergency operations. Details of this effort can be found in S1 Appendix.

### Data sources and analysis

The study period extended from the local epidemic onset in February 2020 through November 2021, prior to the appearance of King County’s 1^st^ Omicron cases. Daily COVID-19 cases, hospitalizations, deaths and test numbers for King County as a whole and by zip code were downloaded from the King County COVID-19 dashboard (7). Population data for King County and Vashon are from the April, 2020 US Census. Zip code level population data are the average of two zip code tabulation area estimates from the WA Office of Financial Management (8) and Cubit (a commercial data vendor providing access to US Census information). The Asset Limited, Income Constrained, and Employed (ALICE) metric, a measure of the working poor, was obtained from United Way (9).

We computed unadjusted cumulative case rates normalized per 1,000 population for Vashon and King County by age and race/ethnicity and compared them by Pearson’s χ^2^ tests. PHSKC provided age, race and gender for all Vashon cases where it was known. Age and gender data for King County and Vashon COVID-19 cases were essentially complete, but 6% of King County cases and 7% of Vashon cases were missing race and ethnicity. Black, Native American and Hawaiian/Pacific Islanders collectively make up less than 1% of the Vashon population and 2% of cases, making accurate assessment of case rates in these populations difficult and posing a risk to confidentiality. However, because these groups are at significant risk from COVID, it was nonetheless important to evaluate the collective risk of these populations. To do this, Vashon’s minority communities were combined into a single Black/Indigenous American/Pacific Islander (B/IA/PI) demographic for comparison with King County and other Vashon populations.

To investigate whether Vashon’s low case rate is an outlier in King County, we developed a multiple linear regression model for cumulative COVID-19 case rates in the 77 King County zip codes with an estimated population greater than 1,000 and that performed more than 500 tests/K population during the study period. To construct the model, we considered a variety of age, race/ethnicity, wealth and educational metrics that might logically be related to COVID-19 case rates. The metric in each of these four categories with the highest R^2^ value in a simple linear regression of COVID-19 rates was then combined into a multiple linear regression model. The 4 metrics employed are the fraction of: 1) population <30 years of age; 2) population that is White or Asian (which have similar low COVID-19 rates in King County); 3) adult population with a Bachelor’s degree; and 4) households below the ALICE threshold (S3 Fig). We used multiple linear regression with heteroscedacticity-consistent (HC3) robust standard errors (10) because analyses showed modest deviation from normality and heteroscedacticity (11-13) (p<0.05). We calculated 95% prediction intervals from the multivariable model using robust standard errors of the point prediction for each observation. Multiple linear regression models of hospitalization and death rates in King County were developed using the same variables and statistical methods.

To directly evaluate the effectiveness of VMRC’s COVID-19 response program we examined the timeliness of VMRC’s vaccination campaign and its case investigation/contact tracing effort. We compared the number of vaccine doses administered over time in King County and Vashon, normalized for total population, using a Kolmogorov–Smirnov two-sample test to compare the two cumulative distributions (14). We chose this metric because it reflects vaccination of the total at-risk population and it is independent of changing eligibility requirements during the study period. To extend this analysis, we similarly compared vaccination of Vashon to the 6 King County zip codes predicted by the regression model to have the most similar case rates, and the 6 zip codes with the lowest observed case rates. We also calculated the time required to reach milestones of 1400-1600 doses/K population, approximating 2 doses for 70-80% of population.

To assess population mobility during the study period, we obtained King County highway and Vashon ferry traffic data from the Washington State Department of Transportation. Using 2019 as a baseline, we compared the fractional quarterly reduction in combined vehicle traffic on Vashon’s three ferries to the reduction in vehicle counts from 21 King County traffic recorders for which complete data were available. To determine the absolute magnitude of mixing between Vashon and surrounding communities, we also calculated the daily number of round trips to and from Vashon (drivers plus passengers and foot-traffic) and normalized it for Vashon’s population.

The study protocol was reviewed and approved by the Human Subjects Protection Program at the University of California, San Francisco (#22-36518). Statistical comparisons were made with the Stata 17 software package (StataCorp LLC, College Station, TX). P-values <0.05 were considered statistically significant for all tests. Data from this study have been submitted to Dryad and are available without restriction upon publication (15). The manuscript was prepared in accordance with STROBE guidelines (16).

## Results

Weekly counts of SARS-CoV2 cases/100,000 population for the study period are shown in Fig 1. As was the case in much of rural America (17), the pandemic was delayed in reaching Vashon, but COVID-19 cases surged in November 2020, reaching 95% of the rate in King County as a whole, and again in October 2021 when Vashon’s rate exceeded the county rate. More than half of all Vashon’s cases observed during the study period occurred during these spikes. However, unlike many other rural communities and King County as a whole, Vashon’s case rate quickly declined producing a cumulative rate 71% lower than that of King County overall (22.0 vs 76.8 cases/1K).

**Fig 1.**
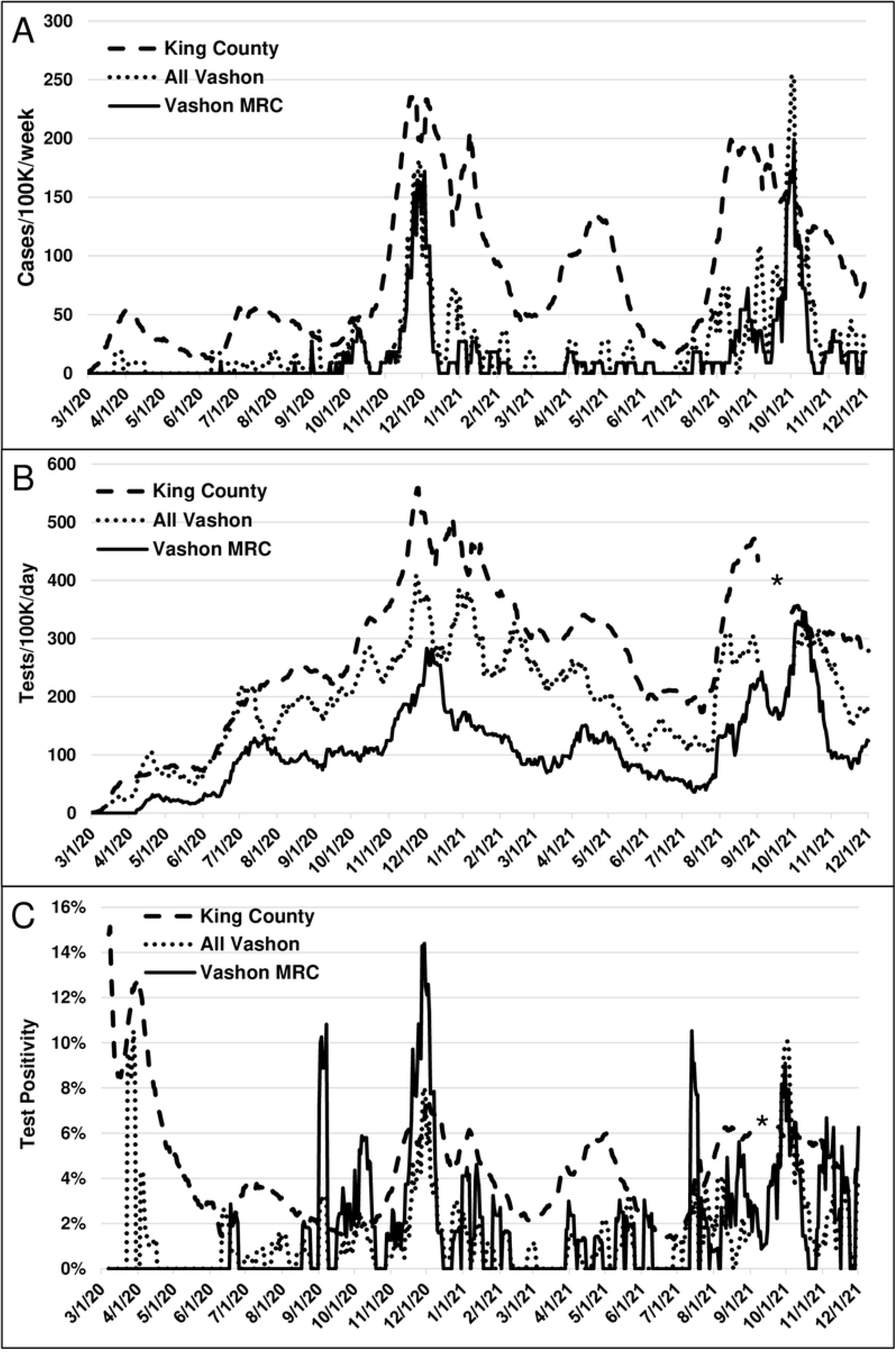
Chronology of SARS-CoV2 cases, tests and test positivity rate for King County and Vashon. A) Cases are presented as a rolling 7-day average normalized per 100K population. Vashon incidence rates approached King County’s during the Beta surge in winter 2020 and exceeded it during the Delta surge in winter 2021. B) Tests are presented as a 14-day rolling average. VMRC performed 41% of all Vashon tests but identified 65% of all Vashon cases. * Indicates a 2-week gap in PHSKC test reporting. C) Despite its low case rate, Vashon’s test positivity rate often approached or exceeded the King County rate- a result of effective contact tracing.

On average 1.5% of Vashon residents were tested weekly, a rate 73% of the King County rate (Fig 1B). Notably, VMRC performed 42% of all tests of Vashon residents, yet identified 65% (157/243) of Vashon COVID-19 cases. Vashon’s cumulative 1.8% overall test positivity rate compares favorably with King County’s 4.5% rate VMRC’s peak test positivity rate exceeded King County’s during local surges (Fig 1C), likely as a result of VMRC’s contact tracing activities described below.

Because Vashon is older (median age 54 vs 37 years) and less racially diverse (86% vs 64% white) than King County as a whole, we compared unadjusted cumulative case rates for Vashon and King County by age and race/ethnicity (Fig 2). Vashon’s case rates were lower across all age groups (Fig 2A, p<.01). Importantly, the greatest difference in case rates were in those older than age 50 for whom COVID-19 poses the greatest risk (1). Vashon’s cumulative case rate was just 20% that of King County in this age group (12 vs 61 cases/K). We calculated an expected case rate for Vashon by applying King County rates for each age group to Vashon’s age distribution. The Vashon case rate predicted from its age demographics is 91% of the observed King County rate.

**Fig 2.**
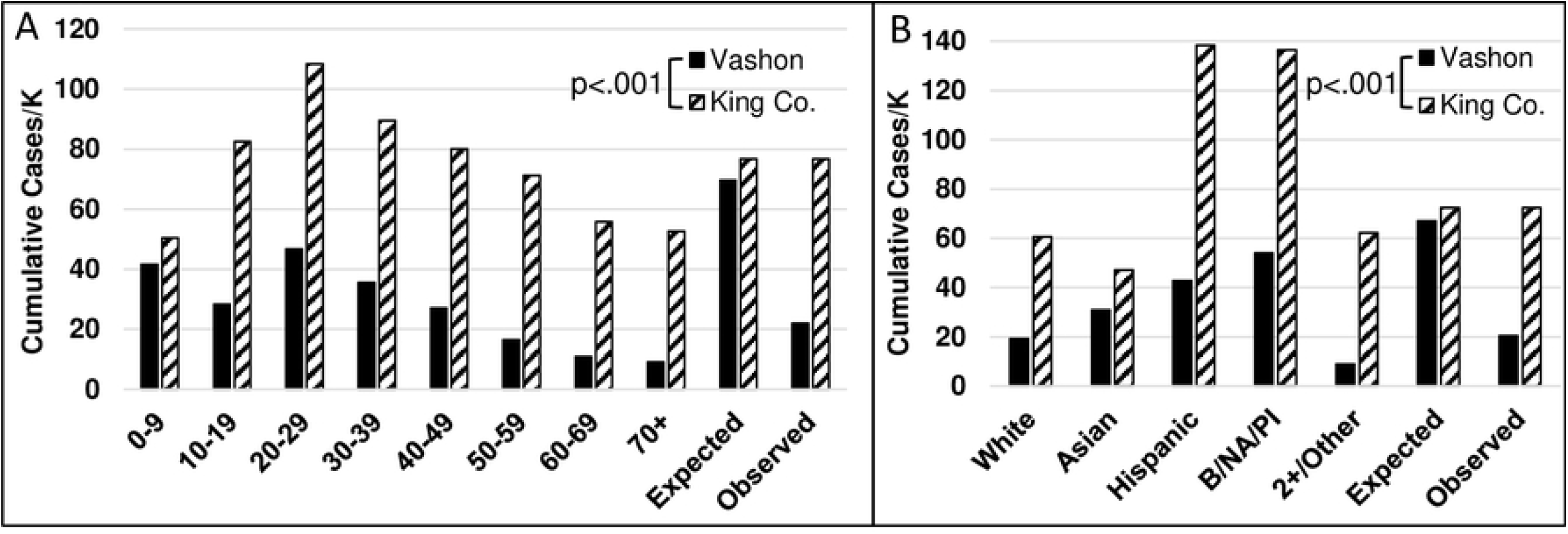
Unadjusted cumulative COVID-19 case rates by age and race/ethnicity. Vashon case rates were lower across all age brackets (panel A, p<.01) and race/ethnicities (panel B, p<.01). Expected rates for Vashon were calculated by applying King County rates for each group to the Vashon population. A single rate was calculated for Black, Native American/Alaska Native, and Pacific/Hawaiian Islanders because these groups collectively represent <1% of the Vashon population.

We performed a similar analysis for race and ethnicity (Fig 2B). Vashon case rates were again lower than King County rates for all groups (p<.01). When King County case rates were applied to Vashon’s demography, the predicted Vashon rate is 93% of the King County rate and more than 3 times Vashon’s actual rate. It is notable that Vashon’s non-white/non-Asian communities, while protected from COVID-19 compared with King County overall, still had cumulative case rates 2-3 times higher than Vashon’s white and Asian communities just as they did throughout King County and the nation (7,18,19).

To extend the demographic analysis, we developed a multiple linear regression model of cumulative COVID-19 rates in King County predicted by age, race/ethnicity, educational attainment and wealth. Components of this model are the fractions of:

- population age <30 years- the age groups with the highest King County rates;
- population that are white or Asian- groups with the lowest King County rates;
- adult population with a Bachelor’s degree; and
- households meeting ALICE criteria (9)- a measure of the working poor.

Predicted and observed cumulative COVID-19 rates for each zip code are shown in Fig 3, ranked by predicted value. Cumulative COVID-19 cases varied by 7-fold across King County zip codes and the regression model explains 88% of the variation. Vashon is a significant outlier with an observed rate 38% of predicted (p<.05) and 35% lower than any other observed rate in King County. No King County zip code was predicted to have a cumulative case rate as low as Vashon’s observed rate. In absolute terms, this translates to 407 fewer observed Vashon cases than predicted from the demographic model.

**Fig 3.**
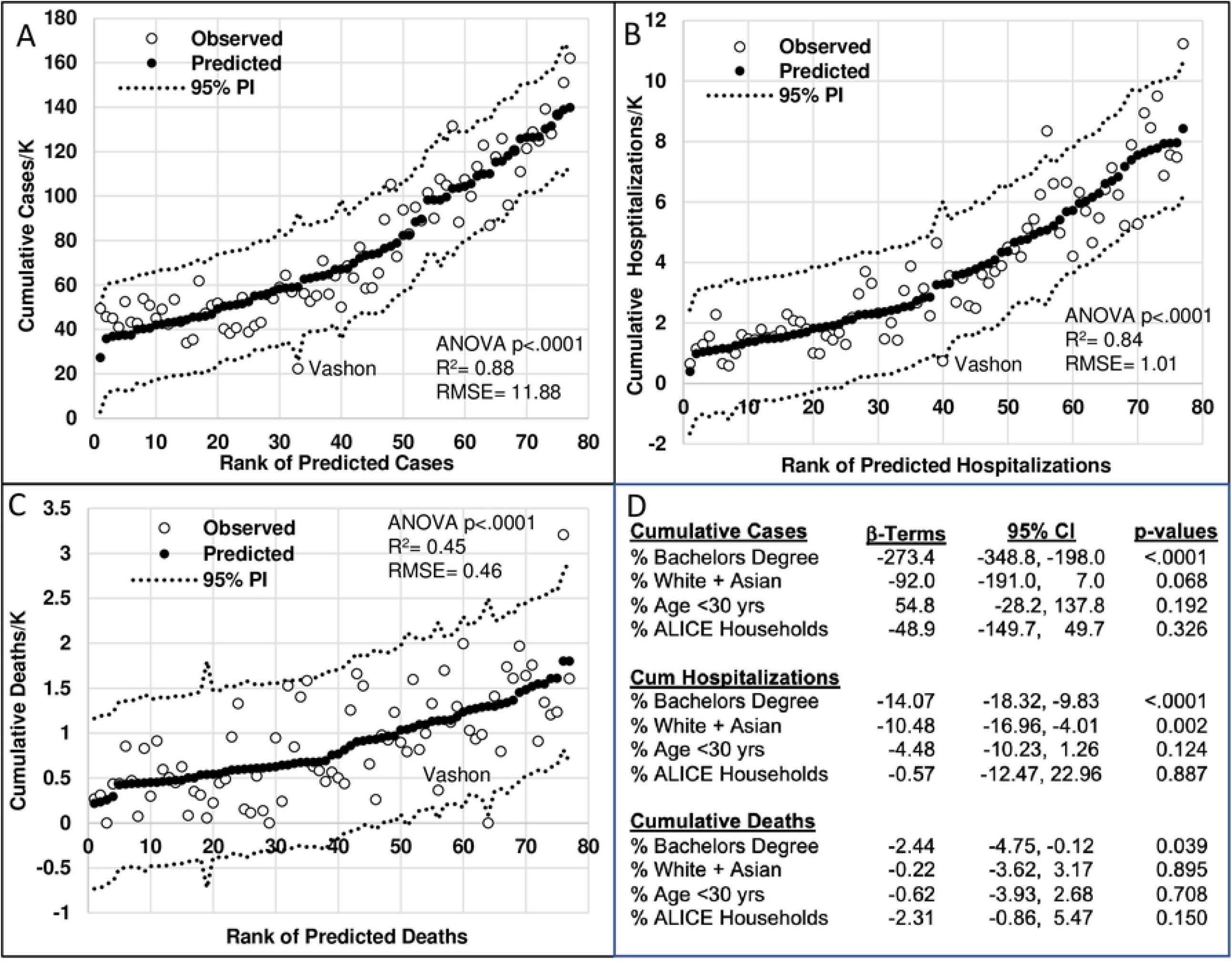
Models of Cumulative COVID-19 Rates in King County, WA. Zip code-aggregated metrics of age, race/ethnicity, education and wealth were used to construct multiple linear regression models of cumulative COVID-19 case rates (A), hospitalizations (B) and deaths (C). Closed circles show predicted case rates in rank order and open circles are observed case rates. Vashon is the only zip code with a case rate below the 95% prediction interval. Beta coefficients, confidence intervals and p-values for independent variables are shown in panel D.

We also modeled cumulative King County hospitalization (Fig 3B) and death rates (Fig 3C). Hospitalizations closely paralleled the model of cases with a similar adjusted R^2^ value, β-coefficients in the same relative order, and Vashon’s predicted rate 4.5 times higher than observed (p=.07). Modeling of death rates showed more scatter with an R^2^ value of 0.50, in part due to the relatively small number of deaths in many zip codes. While not achieving statistical significance, Vashon predictions translate to 28 fewer hospitalizations and 9 fewer deaths than predicted from Vashon demographics. Interestingly, educational attainment is the dominant variable in all three models and the only variable that was significant in each (Fig 3D). Age was a surprisingly poor predictor of COVID-19 cases, hospitalizations and deaths in King County.

Despite its history of vaccine hesitancy (3), Vashon was vaccinated at a significantly accelerated pace compared to King County overall (Fig 4, Kolmogorov–Smirnov, p<0.01). Notably VMRC administered 75% of all doses delivered to Vashon residents. Vashon’s advanced median age meant more of its population was eligible for vaccination, facilitating an early start that allowed the community to reach 1500 doses/K population (a surrogate for 75% vaccination) 4 months earlier than King County overall (184 vs 311 days). Vashon was also vaccinated significantly faster than zip codes predicted to have similar case counts from the regression model and those with the lowest cumulative case rates (Fig 4B, p<.01), reaching the 1500 doses/K milestone 140 days and 78 days faster than zip codes with similar demographics and lowest cases respectively.

**Fig 4.**
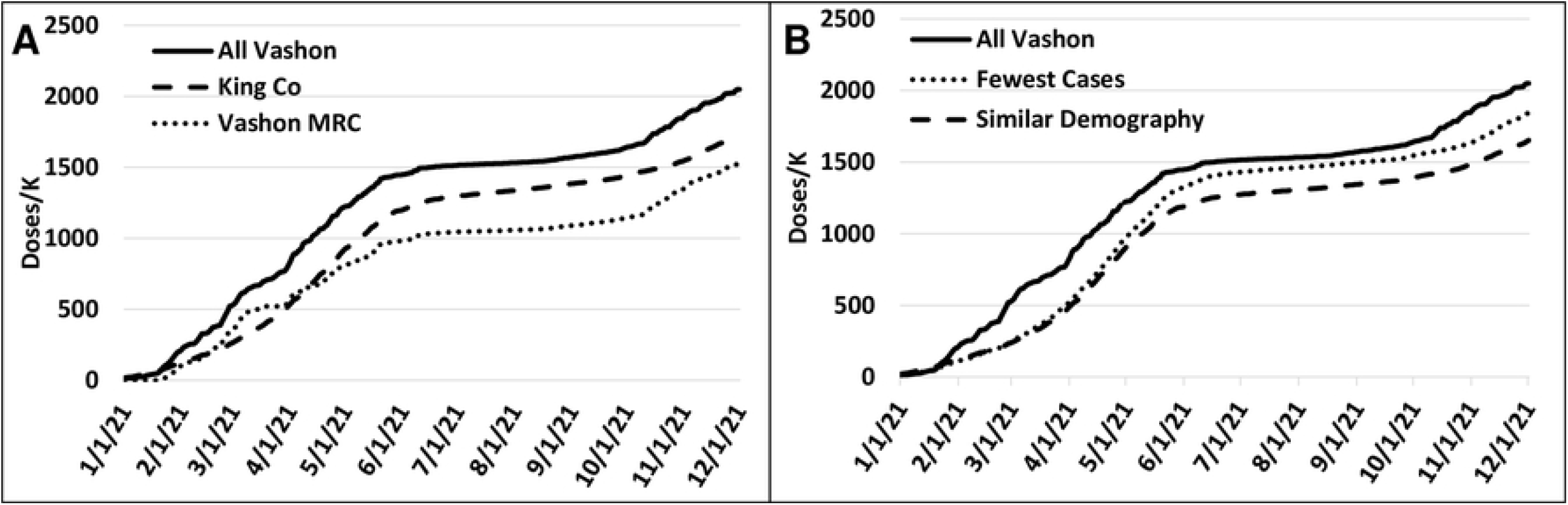
Vaccine accumulation curves for Vashon and King County over time. (A) Cumulative vaccine doses administered for Vashon and King County were normalized for total population. Vashon was vaccinated at an accelerated pace compared with King County (p<.01). VMRC administered 75% of all doses to Vashon residents during the study period. (B) Vashon was also vaccinated faster than the 6 zip codes with the most similar predicted case counts (3 higher and 3 lower) and the 6 zip codes with the lowest cumulative case counts (both p<.01).

To understand the relative impact of vaccination on cumulative COVID-19 rates, we repeated the multiple regression analysis including vaccination rate. As expected, inclusion of vaccination rates in the model resulted in an increase in R^2^ value and decrease in standard error, but the predicted value for Vashon COVID-19 cases remained twice the observed value (S4 Fig), suggesting that vaccination rate only partially explains Vashon’s low COVID-19 rates.

We next examined VMRC’s contact tracing effort. From April 2020 through June 2021 VMRC conducted case investigation and contact tracing for 106 Vashon cases-80 tested by VMRC, 17 referred by PHSKC and 9 self-referred (Fig 5). We were able to interview 97 cases (92% of this cohort) with an average time from test to interview of 1.7 days. Case investigation failed in 8 of 17 PHSKC-referred cases, and just 1 case tested by VMRC. In contrast, due to reporting delays and resource limitations, PHSKC and the WA Department of Public Health *attempted* case investigation of just 75% of Vashon cases through early June, 2021. This cohort could be parsed into 55 index cases and 51 cases among contacts. All contacts named during VMRC interviews were notified, 94% of contacts were tested, and 27% of all contacts tested positive. Notably, 73% of index cases named contacts, but only 28% of cases among contacts did. This reflects the fact that most secondary or tertiary cases were in quarantine before testing and as a result had many fewer contacts (0.6 vs 3.5 contacts per successful interview).

**Fig 5.**
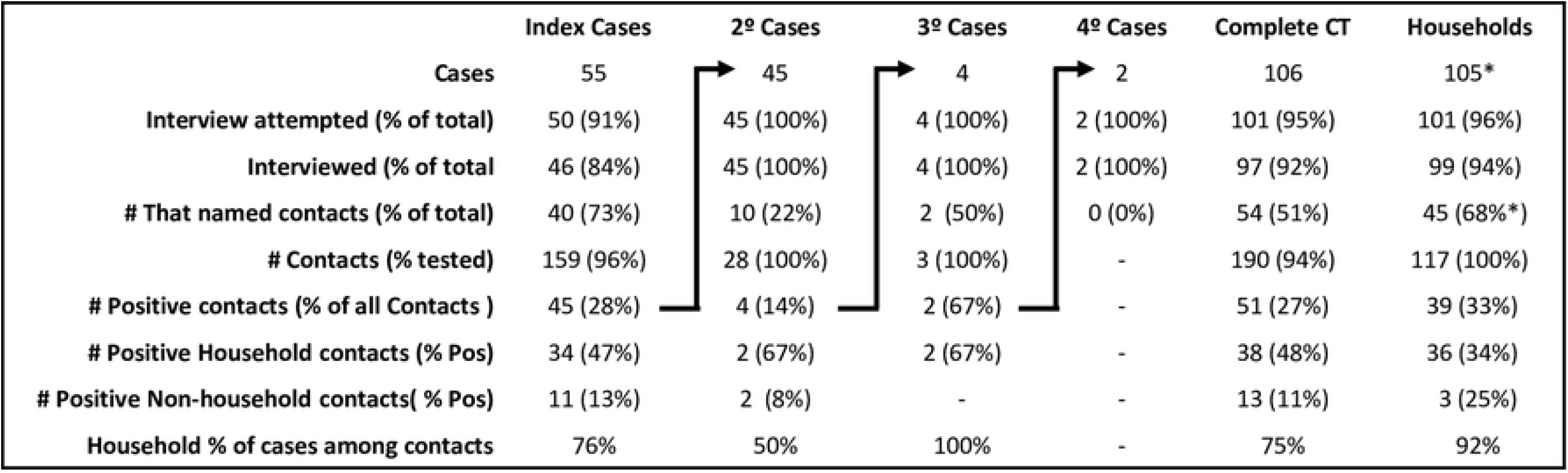
VMRC Case Investigation and Contract Tracing. Cases from April 2020 through June 2021 investigated by VMRC were parsed into 55 index cases and 51 cases among contacts of index, secondary and tertiary cases. The limited number of contacts of non-index cases reflects success of isolation and quarantine. Overall, 31% of contacts tested positive compared with 2% of all Vashon tests. Summary data for this cohort are under the “Complete CT” heading. Household contacts had a 48% positive rate and represented 75% of cases among contacts. Thereafter, a 2^nd^ cohort consisting of 66 index cases focused on household contacts (under “Households” heading). *45 of 66 Household cohort index cases named contacts, similar to the initial cohort.

In this initial cohort we found that household contacts were more than 3 times more likely to test positive than non-household contacts (47% vs 13%). We used this information to prioritize our efforts during streamlining of the volunteer effort in summer of 2021 as VMRC’s contact tracing resources became limiting. From July through November, we focused contact tracing on household contacts while asking individuals to notify potential contacts and have them call VMRC’s hotline for quarantine advice and testing. The experience from July-November 2021 was very similar to the earlier period-VMRC investigated a similar fraction of all Vashon cases and 94% of known cases were interviewed (Table 1) with an average test-to-interview time of 1.6 days. As before, half of cases that could not be reached for interview (3/6) were referrals from PHSKC. As expected, the number of named contacts fell from 2.0 to 1.2 per successful interview, and over 90% of secondary cases were in household contacts.

Finally, because Vashon’s geography may have contributed to its low COVID-19 case rate by reducing mobility (20,21), we compared quarterly reductions in vehicle traffic on Vashon ferries and King County highways (Fig 6). Vehicle traffic reductions on Vashon ferries and King County highways were nearly identical throughout the study period (Fig 5B). Because there is essentially no through-traffic on Vashon, we used Vashon ferry passenger data to estimate the absolute size of the population making a daily roundtrip to or from Vashon. While there is no direct comparator for King County overall, the reduction in passenger traffic to and from Vashon mirrored the overall reduction in vehicle traffic across the study period (Fig 5B). Moreover, a population equivalent to 20-35% of Vashon’s population made a daily roundtrip to or from Vashon during the study period, with a mean of 27% (Fig 5B). Travel to and from Vashon appears to have provided ample opportunity for local seeding of COVID-19 as evidenced by the occurrence of Vashon cases in 19 of the 21 months of study.

**Fig 6.**
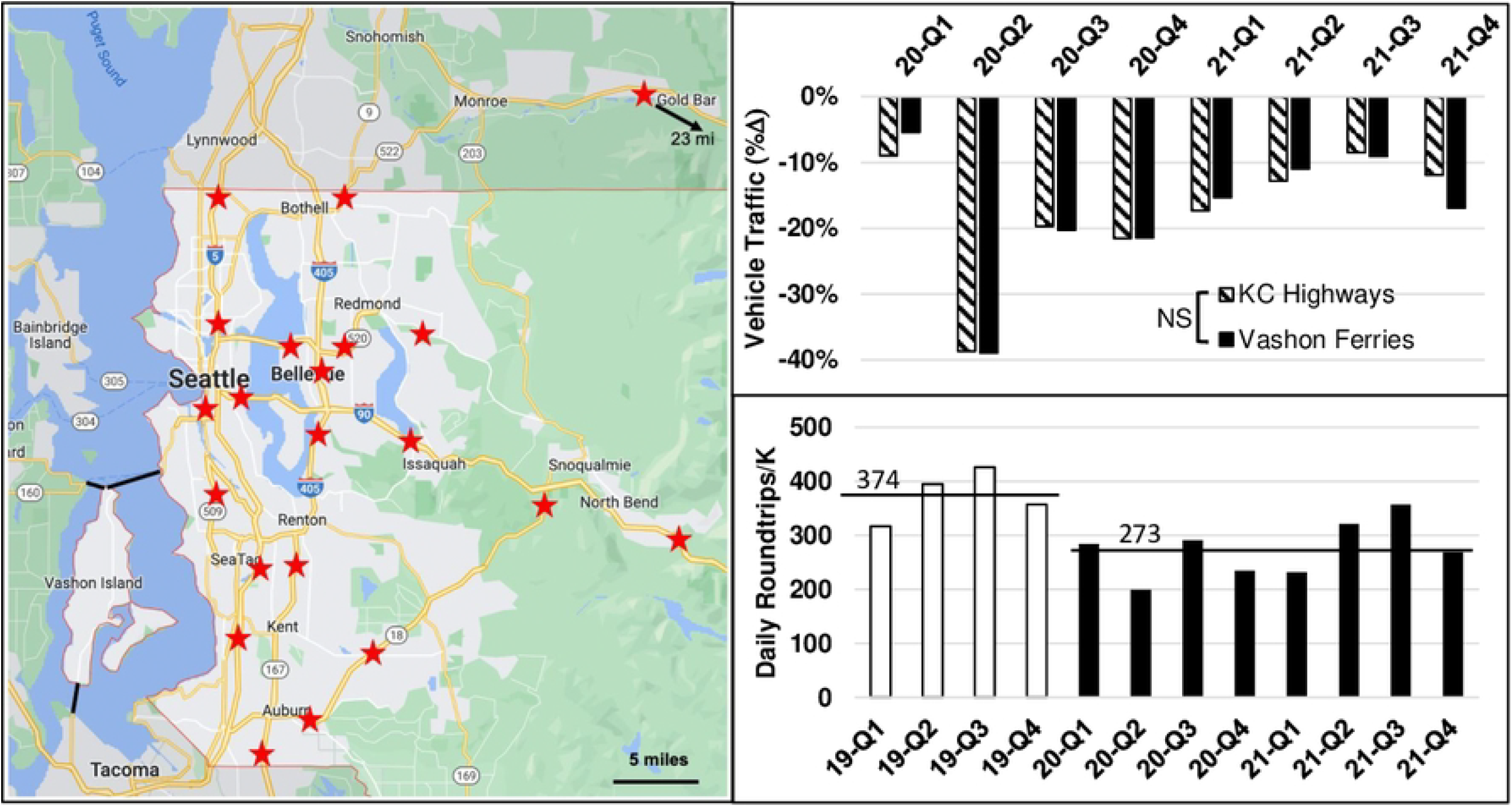
Comparison of pandemic-related mobility in King County and Vashon. A) Map of permanent traffic recorders (red stars) in King County. The three Vashon ferries are shown as black lines from the north and south ends of Vashon. B) Quarterly vehicle traffic for King County and Vashon expressed as the fractional reduction from the same quarter of 2019. Reductions are very similar (p=.93). C) Reduction in daily roundtrip passenger travel to and from Vashon, normalized for population. Open bars are 2019 data and black bars represent the study period. Horizontal lines show averages for 2019 and the study period. Map created with Google Maps.

## Discussion

COVID-19 poses particular risk to rural populations due to an aging population with attendant co-morbitidies and limited access to acute care. Recognizing this risk, VMRC and VashonBePrepared implemented an integrated community-based program of public engagement, testing, contact tracing, vaccination and community support that was associated with a 62% reduction in cumulative COVID-19 cases compared to that predicted from a multivariable linear regression model of COVID-19 in King County, WA. Vashon hospitalizations and deaths were also reduced although these changes did not reach statistical significance. While Vashon’s number of deaths and hospitalizations are small, Vashon ranked in the top quartile of King County zip codes for both case fatality rate (1.7%) and death rate among those hospitalized for COVID-19 (50%), suggesting that the primary benefit of the Vashon COVID-19 response program was reducing COVID-19 transmission.

VMRC’s contact tracing and vaccination programs were particularly effective elements of Vashon’s COVID-19 response. VMRC contact tracing compares very favorably with published US data with respect to speed of case investigation, successful interview rate and contacts notified/tested (22-24). PHSKC did well by national standards (25), but did not attempt to contact 12% of King County cases due to reporting delays and was successful in 80% of attempted interviews, for an overall successful interview rate of 70%. In contrast, VMRC successfully interviewed 93% of all cases reported to it. Vashon conducted interviews in half the time required by PHSKC (25), primarily because test results were directly accessible to the contact tracing team rather than reported to a centralized state-wide system requiring subsequent referral to the county. VMRC delivered test results by telephone rather than email or text, so case investigation could begin immediately. Vashon residents were also much more likely to identify contacts than residents of large metropolitan areas, we speculate because of outreach efforts emphasized the importance of identification and appropriately-timed testing of close contacts.

Despite its history of vaccine hesitancy (3), VMRC working with a local pharmacy was able to rapidly vaccinate the community reaching 1500 doses/K (a proxy for 75% completion of the primary vaccine series) 127 days earlier than King County as a whole and 145 days sooner than zip codes with similar demography. Collectively, our findings do not diminish the effectiveness of PHSKC, but rather suggest that a more decentralized effort can be particularly effective when targeted on rural or other at-risk rural communities.

Our study has limitations. First, the regression model presented was developed to fit local data with the specific goal of understanding whether or not Vashon is a statistical outlier. As such, it may not be generalizable to other areas. Second, we can’t separate the contribution of the several elements of Vashon’s COVID-19 response program to its overall success. While our data show that SARS-CoV2 testing, contact tracing and vaccination were highly effective, integration of COVID-19 response elements is important (26), and we managed that by providing Vashon residents with “one-stop shopping” for COVID-19 concerns. Further, trust is an essential component of successful pandemic responses both globally and locally (27,28) and was likely engendered through VMRC’s public engagement and material support. Third, because Vashon is an island, we cannot completely exclude geographic isolation as a protective factor. Traffic data suggest that is not the case, but may be an imperfect proxy for mobility. Geography likely did contribute to a sense of community that facilitated the Vashon community’s acceptance of COVID-19 control efforts.

Finally, it is not clear whether the VMRC program can be translated to other communities. VMRC clearly benefitted from pre-existing organizational expertise as well as a committed base of registered emergency workers with a broad range of professional experience. Many communities will lack this expertise. However, if provided with appropriate planning, protocols and tools, we believe most communities could carry out this program, especially with the support of a local clinic and their public health department. Indeed, elements of our program were successfully exported and implemented in several rural and tribal Pacific northwest communities.

The activities we describe here are just one example of the under-appreciated effort from the nation’s 800 MRC units, most of which contributed in substantial ways to their local pandemic response. We believe that MRCs can be particularly effective at extending the reach of county public health departments because they are community-based. Expanding the number of MRCs and strengthening their connections with public health departments should be an essential element of ongoing pandemic planning.

## Data Availability

Data have been deposited in Dryad under a paid agreement with UCSF with accession # orcid.org/0000-0002-1971-2273

## Acknowledgements

We thank the members of VMRC’s COVID-19 Steering Committee for tireless support of their community during the pandemic. Committee members are James Bristow, Clayton Olney, Zachary I. Miller, Ina Oppliger, Lydia Aguillar, Bonny S. Olney, Allen de Steiger, Shawn Boeser, Marijke Van Heeswijk and Tyler Young. We thank Julia Hood, Sargis Pagosjans and Kathryn Lau for sharing PHSKC data. We also thank Jay Shendure, and Owatha Tatum for encouraging this work in its early stages and Axel Visel, Nigel Mouncey and Rick Rogers for critical reading of the manuscript. Finally, we thank the Vashon community for accepting the shared responsibility to keep each other safe.

## Supporting Information Captions

**S1 Fig:**
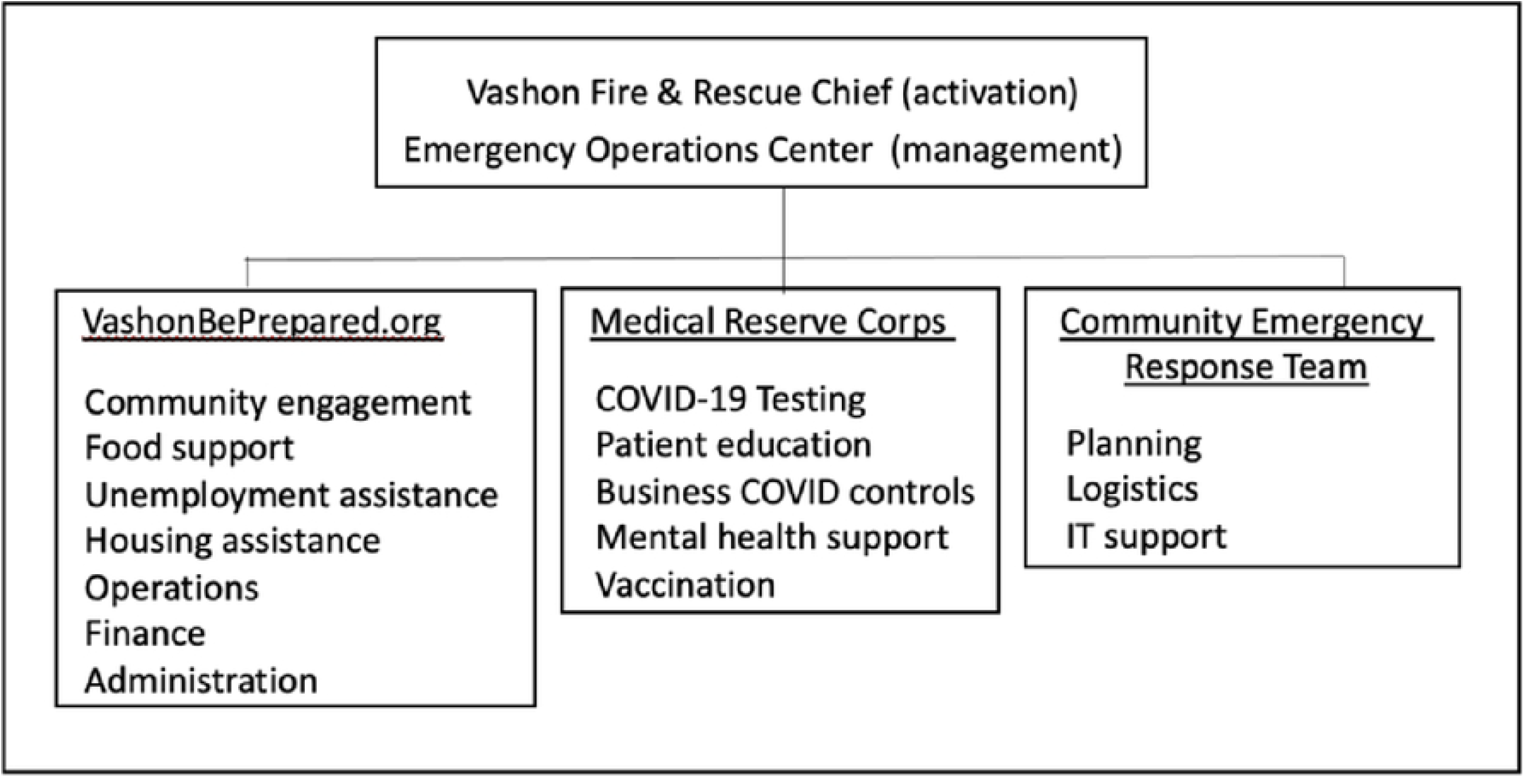
Structure of the Vashon COVID-19 response program. VashonBePrepared.org is a 501c3 organization that houses Vashon MRC and the Community Emergency Response Team for administrative and legal purposes. Following emergency activation by Vashon Fire & Rescue, the Vashon COVID-19 response assumed a typical incident command structure with operational control by an Emergency Operations Center.

The Emergency Operations Center is staffed by volunteers from each of the 3 participating organizations. VashonBePrepared took primary responsibility for community engagement and material support; the MRC had primary responsibility for the health-related activities and the Community Emergency Response Team had responsibility for logistics.

VashonBePrepared raised more than $400,000 for its COVID-19 Relief Fund from hundreds of donors. Because some costs were reimbursed through CARES Act and FEMA funding, the Relief Fund was ultimately able to distribute $546,000 in 4 areas: health, food security, housing security, and economic recovery. The testing and contact tracing effort that is the main thrust of this paper had a monthly cost of $1,200 as tests were largely paid by patient insurance. Uninsured patient tests were covered by the CARES Act or the COVID-19 Relief Fund.

The Relief Fund supported food security through the funding of the Vashon Maury Community Food Bank, the Vashon Senior Center and the Vashon Island School District Nutrition program, resulting in the distribution of more than 25,000 meals and 4,300 bags of groceries. VashonBePrepared, working with the local Chamber of Commerce, provided economic relief by helping 400 residents with applications for unemployment and other state or county benefits. The Relief Fund also provided emergency rent relief and other support to over 400 families needing assistance. Vashon’s Hispanic community received specific attention from these relief efforts.

### Demographics & Testing for COVID-19: Vashon and King County, WA

**S2 Table:** VMRC testing, Vashon and King County demographics. Vashon and King County total populations are from the 2020 US Census, and age and race/ethnicity compositions are from the 2019 American Community Survey estimate. Forty percent of the Vashon population is over age 60-twice the fraction in King County overall. VMRC testing was skewed toward younger age groups. VMRC tests were roughly reflective of Vashon’s racial/ethnic makeup, with slight under-sampling of the Hispanic community and over-sampling of the Black and Hawaiian/Pacific Islander communities.

| **Age** | **VMRC Tests** | **Vashon Population** | **King County Population** |
| --- | --- | --- | --- |
| 0-9 | 7.1% | 7.0% | 11.0% |
| 10-19 | 11.5% | 10.2% | 11.2% |
| 20-29 | 10.2% | 7.4% | 15.1% |
| 30-39 | 13.4% | 8.4% | 17.6% |
| 40-49 | 15.5% | 11.7% | 13.6% |
| 50-59 | 17.6% | 15.9% | 12.3% |
| 60-69 | 14.2% | 21.6% | 10.2% |
| 70+ | 10.6% | 17.9% | 9.0% |
| Total tests or population | 3,566 | 11,095 | 2,269,675 |
| **<u>Race/Ethnicity</u>** |  |  |  |
| White | 85.4% | 84.1% | 54.2% |
| Hispanic | 5.3% | 5.7% | 10.7% |
| Multi-racial | 5.0% | 7.2% | 7.4% |
| Asian | 2.5% | 2.0% | 19.8% |
| Black | 1.1% | 0.7% | 6.5% |
| Hawaiian, Pacific Islander | 0.5% | 0.1% | 0.9% |
| Native Amer/ AK Native | 0.2% | 0.2% | 0.5% |
| Total tests or population | 3,240 | 11,095 | 2,269,675 |

**S2 Fig:**
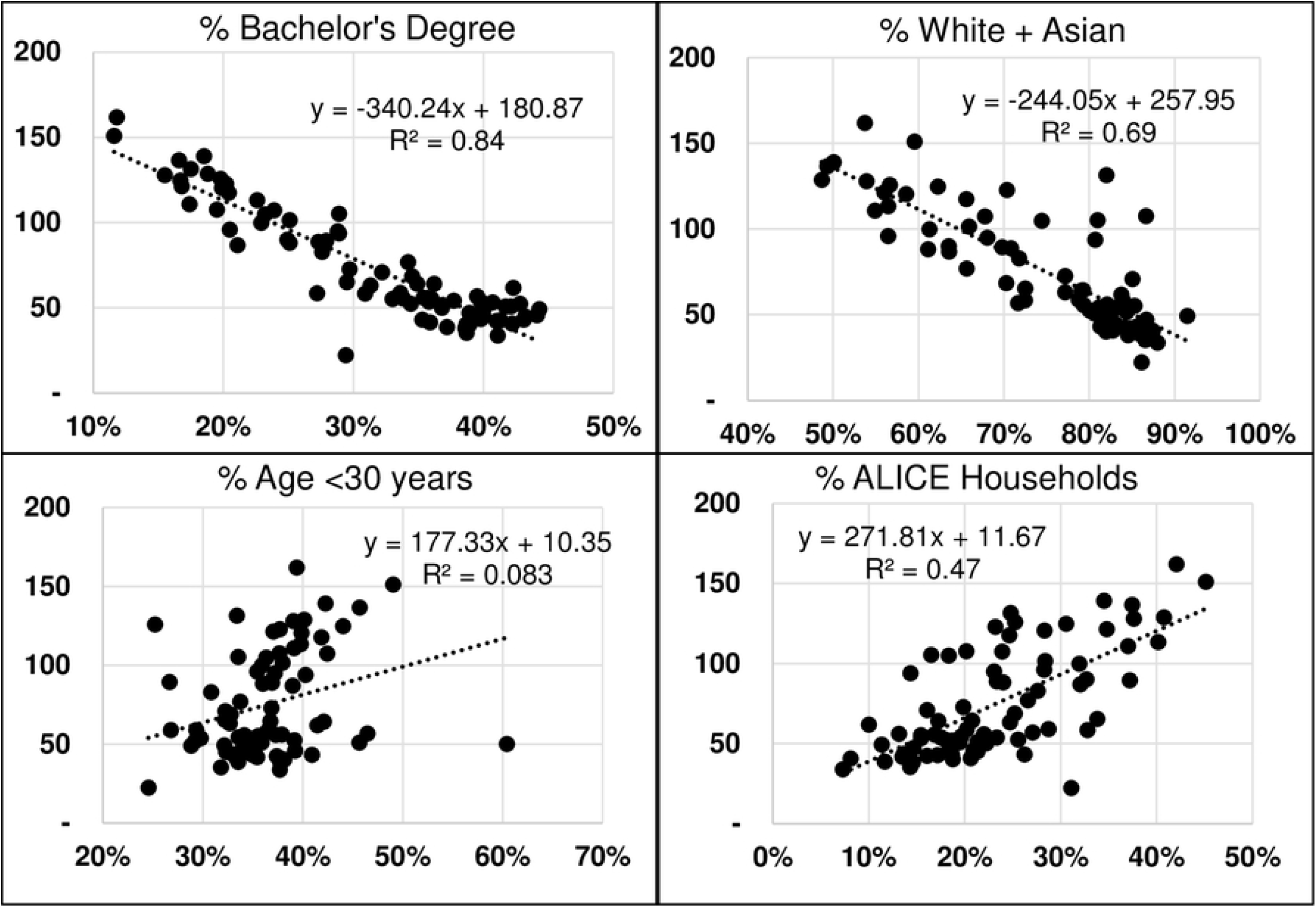
Components of the multiple linear regression model for cases. Ordinary least squares linear regression of COVID-19 cases was carried out against several logical variables of age, race/ethnicity, educational attainment and wealth that might be associated with cumulative COVID-19 rates. Those with the highest R2 value are above. Multiple age groupings were evaluated and while none performed well, age <30 had the highest R^2^ value for cases (above) as well as hospitalizations and deaths (not shown). Total population, population density, and testing rate were also considered for inclusion in the model, but were not correlated with cumulative COVID-19 rates in King County.

**S3 Fig.**
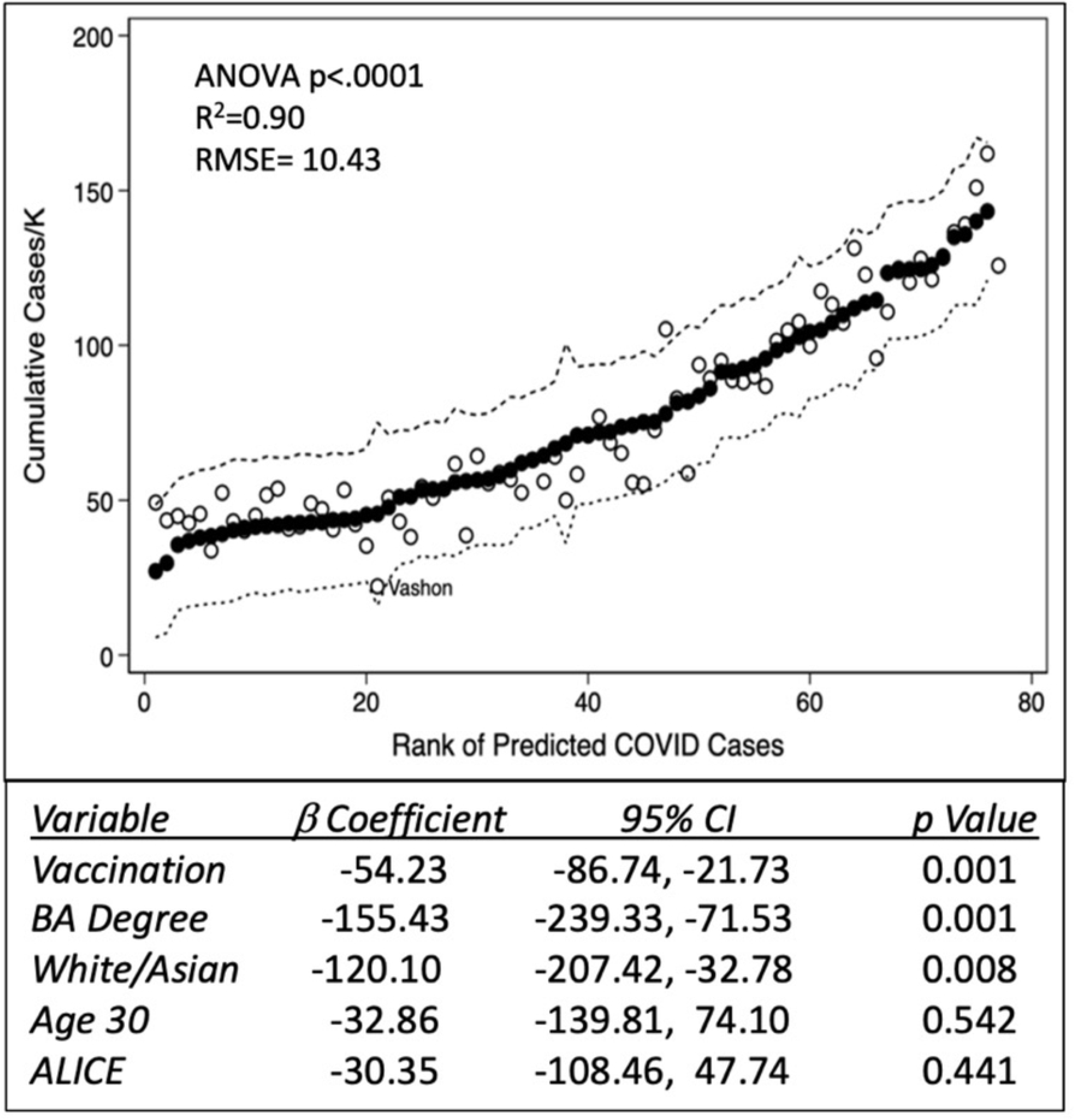
Regression model of King County cumulative COVID-19 cases including vaccination rates. Inclusion of vaccine doses administered/K population improves the R^2^ value and standard error of regression analysis carried out as in Fig 3. Vashon’s predicted rank improves from 32 to 21 and while it is no longer a statistical outlier, its predicted rate remains twice its observed rate.

